# Large language models in simplifying radiological reports: systematic review

**DOI:** 10.1101/2024.01.05.24300884

**Authors:** Yaara Artsi, Vera Sorin, Eli Konen, Benjamin S. Glicksberg, Girish Nadkarni, Eyal Klang

## Abstract

**Objectives:** Simplifying medical information to make it understandable for patients, specifically in the case of radiology reports, is challenging. It requires time and effort from medical personnel. This systematic review focuses on the application of large language models (LLMs) in generating simplified radiological imaging reports, as well as answering patient inquiries regarding radiological procedures.

**Materials and Methods:** The authors searched for studies published up to January 2024. Search terms focused on LLMs generated simplified radiological reports and answers to patient inquiries regarding radiological procedures. MEDLINE was used as a search database.

**Results:** Overall, eight studies published between May 2023 and November 2023 were included. All studies showed that LLMs can produce simplified medical information for patients. Four studies (50%) used GPT-3.5, Two studies (25%) conducted a comparative analysis between GPT-3.5 and GPT-4. One study (12.5%) examined Microsoft Bing. One study (12.5%) utilized GPT-4. Four studies (50%) used LLMs to simplify radiological reports. Four studies (50%) used LLMs to answer patient questions regarding radiological procedures. Only two studies (25%) used patients to evaluate the LLMs output. One study (12.5%) compared their initial prompt with optimized prompt. Five studies (62.5%) showed missing, inaccurate and potentially harmful AI outputs.

**Conclusion:** LLMs can be used to simplify medical imaging reports and procedures, for improved patient comprehension. However, their limitations cannot be ignored. Further study in this field is essential and more conclusive evidence is needed.

## Introduction

Reading, understanding, and interpreting radiographic images, reports and procedures is challenging, even for non-radiologist physicians [1, 2]. The task is many more times difficult for patients who have no medical training [3, 4]. Nowadays, medical imaging is an integral part of the clinical decision making process [5, 6, 7].

According to the patient-centered care approach, patients should be active participants in their care [4, 8]. Although patients have access to their imaging reports, these reports are frequently incomprehensible to the average patient [3, 4]. Using complex medical terminology can create patient anxiety and a perception of exacerbated severity of their condition [9]. The lack of access to simplified information for patients regarding their healthcare highlights a shortcoming of modern healthcare practice.

Large language models (LLMs), such as ChatGPT, can be used to analyze free-text and generate human-like responses to various inquiries [10, 11]. It is possible that this technology might hold the key to bridge the gap between patients and complicated medical imaging reports jargon. However, the possibility of patients seeking clarification from AI poses risks. The accuracy and credibility of such models is still up for debate and can potentially misinform patients’ diagnoses and outcomes [12].

The aim of this study is to systematically review the literature on applications of LLMs for patient education and simplification of radiological reports and procedures.

## Methods

### Literature search

For this retrospective review, we conducted a search to identify studies describing LLMs’ applications for patient education. We searched PubMed/MEDLINE for papers published not earlier than 2023 up to January 2024. The following keywords were used with Boolean operators AND/OR: *large language models, ChatGPT, openAI, patient, education*.

We checked the references list of selected publications for more relevant papers. Sections as ‘Similar Articles’ (e.g., PubMed) were also inspected for possible additional articles.

Our study followed the Preferred Reporting Items for Systematic Reviews and Meta Analyses (PRISMA) guidelines. The study is registered with PROSPERO

### Inclusion and exclusion process

Publications resulting from the search were initially assessed by one author (YA) for relevant titles and abstracts. Next, full-text papers underwent an independent evaluation by two authors (EK and VS).

We included full length studies describing LLMs application for patient education focusing on radiology and imaging reports. We excluded papers published before 2023, non-English papers and non-original studies (**Figure 1**).

Discrepancies were discussed and resolved to achieve a consensus. Risk of bias and applicability were evaluated using the tailored QUADAS-2 tool (**Figure 2**).

## Results

### Study selection and characteristics

The initial literature search resulted in 729 articles. Eight studies met our inclusion criteria (**Figure 1**). Six studies were retrospective. One study is cross-sectional (descriptive). Majority of the studies used ChatGPT (versions 3.5 or 4) as an AI model of choice, one study used Microsoft’s Bing. The prompts were phrased differently in each study. One study conducted a comparison between initial and optimized prompt. Two studies involved patients’ evaluation of the simplified reports (**Figure 3**.).

### Descriptive summary of results

Lyu et al. [13] collected a total of 138 imaging reports, 76 chest CT and 62 brain MRI (**Table 2**). All reports were anonymized. GPT-3.5 was provided with three initial prompts requesting simplification of the reports (**Table 7**). The evaluation focused on three aspects: overall score, completeness, and correctness. The number of places with missing information and with incorrect information was recorded as well. Two radiologists evaluated the results using a 5-point system and word count. Patients did not evaluate the simplified reports. For the chest CT reports, 85.5% of the translated results (53 of 62) were shorter than the corresponding original reports. The overall length reduction was 26.7%. In brain MRI radiology reports, 72.4% of the translated results (55 of 76) contained fewer words than the corresponding original reports. The overall length reduction was 21.1% (**Table 3**).

Negative performance of ChatGPT was documented as well, showing instances of inaccurate, missing or incorrect information (**Table 5**). They’ve also compared GPT-3.5 to GPT-4, with GPT-4 outperforming in all aspects. Lastly, they tested the difference between the original prompt and an optimized prompt. The overall quality of translation increased from 55.2% to 77.2%, and the measures on information that were completely omitted, partially translated, and misinterpreted were reduced to 9.2%, 13.6%, and 0%, respectively (**Table 4**).

Li et al. [14] randomly sampled 100 radiographs (XR), 100 ultrasound (US), 100 CT, and 100 MRI radiology reports (**Table 2**). They prompted GPT-3.5 for simplified reports. Mean report length, Flesch reading ease score (FRES), and Flesch-Kincaid reading level (FKRL) were calculated for each original report and GPT-3.5 simplified output (**Table 3**). Patients did not evaluate the simplified reports. Negative GPT-3.5 performance was not detailed. Following simplification by GPT-3.5, all reports had a FKRL <8.5 and 77/100 (77%) of XR, 76/100 (76%) of US, 65/100 (65%) of CT, and 58/100 (58%) of MRI reports had a FKRL <6.5 (**Table 3**)

Grewal et al. [15] tested GPT-4 application in radiology across several fields, including patient education. GPT-4 generated patient-oriented explanations of radiological findings, and assisted in patient inquiries. Patients did not evaluate the simplified reports. Negative GPT-4 performance was not detailed (**Table 3**).

Kuckelman et al. [16] selected three common radiologic examinations and procedures: CT, MRI, and bone biopsy. Ten patient questions for each type of examination or procedure were compiled (**Table 2**). This is the only study that utilized Microsoft Bing. The questions were asked on three different chatbot settings in two trials, for a total of 360 reviews. Attending radiologist and a fourth-year medical student rated the responses independently for accuracy and completeness on a 1–3 scale. They used radiologyinfo.org, an accepted online resource for comparison [17]. The Fleisch-Kincaid level of readability was also examined. Overall, 336 (93%) ratings were “entirely correct”, and 235 (65%) ratings were “complete”. No responses were rated as “inaccurate/incomplete” by either reviewer. The Fleisch-Kincaid level of readability was an eighth-grade level (**Table 3**). Patients did not evaluate the simplified reports. Negative Microsoft Bing performance included missing details about bone biopsy procedure (**Table 5**).

Jeblick et al. [18] wrote three fictitious radiology reports, for knee MRI, head MRI and whole-body CT. The reports were simplified by prompting GPT-3.5. They generated 15 different simplified reports per original report, 45 in total (**Table 2**). Many different prompt designs were tried. Radiologists evaluated the quality of the simplified reports using a 5-point Likert scale in three categories: factual correctness, completeness and potential harm. All simplified reports were factually correct and complete. For quality criteria 75% rated “Agree” (**Table 3**). Negative GPT-3.5 performance included incorrect text passages in 23 simplified reports (51%), missing relevant information for 10 simplified reports (22%) and potentially harmful conclusions for 16 simplified reports (36%) (**Table 5**). Patients did not evaluate the simplified reports.

Scheschenja et al.[19] compiled 133 questions related to three specific interventional radiology procedures (Port implantation, percutaneous transluminal angioplasty and transcatheter arterial chemoembolization). They assessed both GPT-3.5 and GPT-4 responses. The chatbot was primed to respond to specific inquiries (**Table 2**). Grading was performed using a 5-point Likert scale. For “completely correct” GPT-3.5 scored 30.8%lJwhile GPT-4 scored 35.3%. GPT-4 was found to give significantly more accurate responses than GPT-3 (*p*lJ=lJ0.043) (**Table 3**). Negative performance included “mostly incorrect” responses in 5.3% of instances for GPT-3. For GPT-4 just 2.3%. No response was identified as potentially harmful (**Table 5**).

Gordon et al. [20] assessed GPT-3.5 for accuracy, relevance and readability in answering patient imaging-related questions. They compiled 22 imaging-related questions (**Table 2**). The categories for the questions included: safety, the radiology report, the procedure, preparation before imaging, meaning of terms and medical staff. Questions were posed to ChatGPT with and without a prompt. Four board-certified radiologists evaluated the answers for accuracy, consistency and relevance. Two patients also reviewed the responses. Readability was assessed by Flesch Kincaid Grade Level (FKGL). For accuracy GPT-3.5 scored 87% (229/264). Consistency of the responses was 86% (76/88). Nearly all responses 99% (261/264) were partially relevant for both prompt and non-prompt questions. The average FKGL was high at 13.6. When provided with a prompt, GPT-3.5 performed better in all parameters (**Table 3**). Negative GPT-4 performance was not detailed.

Schmidt et al. [21] evaluated the ability of GPT-3.5 for simplifying radiological MRI findings. They created five versions of a simplified radiological report using ChatGPT 3.5 (**Table 2**). They created different prompts until one prompt was selected for the best outcomes (**Table 4**). They asked GPT-3.5 for varying levels of complexity: simple, moderate and complex. Two orthopedic surgeons and two radiologists evaluated the reports for quality, completeness and comprehensibility using a questionnaire. All simplified reports were evaluated by 20 patients. They used a patient-specific questionnaire for comprehension and simplification. The simplified radiology reports were factually correct regardless of complexity. The majority of participants indicated “Agree” with respect to the simplicity and comprehensibility (**Table 3**). Negative performance included missing 53.8% (7/13) or incorrect 23% (3/13) information across all simplified findings. For potentially harmful conclusions the simplified reports misinterpreted crucial information 6 times. An incorrect need for therapy was indicated two times, and degeneration was interpreted as injury once (**Table 5**). In addition, patient evaluation showed that while they knew what the text was about, the majority responded that the simplified text did not inform them as well as a doctor.

Some studies presented examples for different prompts they used, as well as examples for the simplification process done by ChatGPT. We included examples from each study of simplification and prompt generation, presented in **Table 6** and **Table 7**, respectively.

## Discussion

In this review we examined LLMs capability to simplify radiological reports and procedures, enhancing patient comprehension and education. All studies reviewed demonstrated LLMs capability in generating simplified, understandable radiological reports.

In the past, radiology was considered a paraclinical field [22]. Radiology reports were written for referring physicians and healthcare providers. Nowadays, radiology emerges to be more clinical and patient centered [23]. Patients can access their imaging reports, but the reports readability is still complex and incomprehensible [24].

Making medicine approachable to patients is a formidable challenge to the medical community. Doctors often use complicated medical terms that patients have trouble understanding [25]. Patients’ misperception of medical jargon can lead to confusion, stress and overtreatment [26, 27]. The application of advanced AI to explain and simplify imaging reports and procedures could be a step toward accessible and approachable medicine.

### LLMs benefits

One of the most limited resources for a medical doctor is time [28, 29]. Working long hours, with many tasks and responsibilities, leaves little time to address patients’ inquiries and concerns [30]. AI continues to evolve, becoming more integrated in various medical applications [31]. AI performance is fast and efficient [32]. Utilizing LLMs chatbots for patient education can save time for both the overworked physician and the patient waiting for answers [33].

Another important advantage is the chatbot’s ability to simplify complicated medical texts into plain language [34]. Gotlieb et al. showed that several common medical phrases are often misunderstood. The interpreted meaning is frequently the exact opposite of what is intended [35]. LLMs ability to make medical terms understandable to patients can alleviate patient concern and anxiety [36].

Lastly, ChatGPT reached over 100 million users in only 2 months [37]. This rapid adoption highlights its potential role in improving accessibility of medical information to patients seeking answers. In every study we reviewed, LLMs significantly improved clarity and simplicity of radiological reports. These models may provide the solution to the knowledge gap between patients and their medical information.

### LLMs drawbacks

When patients rely on LLMs for simplifying their medical imaging reports, they also need to be certain of the medical accuracy. A known limitation for LLMs is called “hallucination”. This occurs when generative AI misinterprets the given prompt, resulting in outputs that lack logical consistency. When relying on AI for accurate medical information, this phenomenon is unacceptable [38]. Also, LLMs can often misinterpret clinical findings [39]. For example, some of the studies we reviewed presented AI output that mistook benign findings as malignant. This can lead to needless patient anxiety and interfere with the physician ability to reach the correct diagnosis [40]

Another concern is the patient’s medical information safety. Medical imaging reports hold private medical information, and can often be the target of cyber-attacks [41]. Finally, another consideration is the disparity between different age groups in their ability to use technology. When applying LLMs for simplifying patients’ imaging reports, a certain level of technological abilities is needed. Older individuals might find it harder to apply LLMs to obtain readability for their medical reports [42, 43].

It is imperative to take into consideration those significant shortcomings and challenges. LLMs should be used with caution while utilized to simplify important medical information.

### Prompt engineering

For each study we examined the process of crafting the prompts. We noticed a wide range of approaches to writing and designing prompts. Only one study [13] conducted prompt optimization, which significantly improved the LLM’s outputs. This emphasizes the importance and sensitivity of prompts. Prompt-engineering may be a task that requires specific training, so that the prompt is phrased correctly and the quality of the simplified medical report is not impaired.

### Limitations

Our review has several limitations. Due to heterogeneity in study design and data, a meta-analysis was not performed. Only two studies tested the simplified result with patients. Several studies used word count as representation of simplification. However, a shorter text is not always a guarantee for simplification. This was not examined. Only one study conducted prompt optimization and evaluated its outcomes. Two studies were at high risk of bias. One study did not present clear parameters for evaluation of bias. Additional studies will be needed to further solidify the usefulness of LLMs in simplifying radiological reports and clarifying radiologic procedures. Lastly, we limited our search to PubMed/MEDLINE. We did so due to its relevance in biomedical research. We recognize this choice narrows our review’s scope. This might exclude studies from other databases, possibly limiting diverse insights.

## Conclusion

Utilizing LLMs for simplifying medical imaging reports and procedures is feasible. In the majority of the studies we reviewed, LLMs demonstrated promise in their capability to generate accessible medical imaging reports. However, their use warrants cautious, critical evaluation. Awareness of LLMs limitations is needed in order to avoid misuse and harming patients’ diagnosis and treatment. Currently, further research in this field is warranted. Until further advancements are achieved, AI should be used with caution when applying it to simplify medical information for patients.

## Additional information

## Data Availability

All data produced are available online at PubMed

https://pubmed.ncbi.nlm.nih.gov/

## Acknowledgments

None

## Disclaimer

None

