## Supplemental Tables 1-7 for "Large language models in simplifying radiological reports: systematic review"

**Table 1**. A summary of the studies included in the review

| **Study** | **Month** | **Journal** | **Study Design** | **AI tool** |
| --- | --- | --- | --- | --- |
| **Lyu et al.** | **May** | **Visual Computing for Industry,  Biomedicine, and Art** | **Retrospective** | **Chat-GPT 3.5/4** |
| **Li et al.** | **June** | **Clinical Imaging** | **Retrospective** | **Chat-GPT 3.5** |
| **Grewal et al.** | **June** | **Cureus** | **Cross-sectional** | **Chat-GPT 4** |
| **Kuckelman et al.** | **September** | **Academic Radiology** | **Retrospective** | **Microsoft Bing** |
| **Jeblick et.al** | **October** | **European Radiology** | **Prospective** | **Chat-GPT 3.5** |
| **Scheschenja et al.** | **October** | **CardioVascular and Interventional Radiology** | **Retrospective** | **Chat-GPT 3.5/4** |
| **Gordon et al.** | **October** | **JACR** | **Retrospective** | **Chat-GPT 3.5** |
| **Schmidt et al.** | **November** | **Archives of Orthopaedic and Trauma Surgery** | **Retrospective** | **Chat-GPT 3.5** |

**Table 2**. A summary of modalities or procedures and numbers of reports before and after simplification

| **Study** | **Modality or Procedure** | **No. Of Original Reports or Patient Inquiry** | **No. Of Simplified Reports or Answers To Patients** |
| --- | --- | --- | --- |
| **Lyu et al.** | **CT  MRI** | **138** | **138** |
| **Li et al.** | **XR  US  CT  MRI** | **400** | **400** |
| **Grewal et al.** | **N/A** | **N/A** | **N/A** |
| **Kuckelman et al.** | **CT  MRI  Bone biopsy** | **30** | **360** |
| **Jeblick et.al** | **MRI / CT** | **3** | **45** |
| **Scheschenja et al.** | **Port implantation  PTA  TACE** | **133** | **133** |
| **Gordon et al.** | **General** | **22** | **132** |
| **Schmidt et al.** | **MRI** | **3** | **15** |

**Table 3**. Presents overall evaluation of LLMs output and performance

| **Study** | **Methods For Evaluation** | **Professional Evaluation after simplification** | **Patient Evaluation after simplification** | **Performance Score** |
| --- | --- | --- | --- | --- |
| **Lyu et al.** | **● 5-point scoring system ● Word count** | **Two Radiologist** | **N/A** | **● 5-point scoring system: 76% best score (5 of 5) - CT 32% best score (5 of 5) - MRI ● Word count: 85.5% shorter (53 of 62) - CT 72.4% shorter (55 of 76) - MRI** |
| **Li et al.** | **● Flesch-Kincaid reading level (FKRL) ● Flesch reading ease score (FRES) ● Word count** | **N/A** | **N/A** | **● FKRL: All reports < 8.5 Reports with FKRL < 6.5 77/100 (77%) of XR 76/100 (76%) of US 65/100 (65%) of CT 58/100 (58%) of MRI ● FRES: 83.5 ± 5.6 ● Word count: Avarage of all combined reports  Original 164 ± 117  Simplified 103 ± 36 Ddifference −61 ± 105  (p value of < 0.01)** |
| **Grewal et al.** | **N/A** | **Expert radiologist** | **N/A** | **N/A** |
| **Kuckelman et al.** | **● 1-3 scale comparison to radiologyinfo.org ● Flesch-Kincaid reading level (FKRL)** | **Attending radiologist Furth-year medical  student** | **N/A** | **● 1-3 scale: 336 (93%) ratings - “entirely correct” 235 (65%) ratings - “complete” ● Flesch-Kincaid reading level (FKRL): at 8th grade level** |
| **Jeblick et.al** | **● Likert scale Questionnaire** | **Fifteen radiologists varying levels of experience** | **N/A** | **● Likert scale:  75% “Agree” or “Strongly agree” with simplified report findings** |
| **Scheschenja et al.** | **● Likert scale Questionnaire** | **Two board-certified radiologists** | **N/A** | **● Likert scale: Chat-GPT 3.5: 30.8% completely correct Chat-GPT 4:   35.3% completely correct** |
| **Gordon et al.** | **● RadiologyInfo Website ● Flesch Kincaid grade level (FKGL) ● 1-3 Scale for patients** | **Four board-certified radiologists** | **Two patient advocates** | **● RadiologyInfo Website comperison: Accurate 79% - 96% Safety 71%. Relevant 67% - 80% ● Flesch Kincaid grade level (FKGL): 13.0 None were at or below the 8th-grade reading level ● 1-3 scale: 97% (128 out of 132) relevant and helpful** |
| **Schmidt et al.** | **● Likert scale Questionnaire** | **Two orthopedic surgeons Two radiologists at least 5 years of professional experience** | **20 Patients** | **● Likert scale: Relevant and comprehensible - “Agree” Simplicity and sentence structure - “Agree” For Patients: Better at knowing what the text was about (p = 0.001) Correct conclusions from simplified report (p = 0.013829)** |

**Table 4**. Presents Prompt generation process

| **Study** | **No. Prompts Generated** | **Initial Prompt Performance** | **Optimize Prompt Performance** |
| --- | --- | --- | --- |
| **Lyu et al.** | **5** | **0.552** | **0.772** |
| **Li et al.** | **1** | **N\A** | **N\A** |
| **Grewal et al.** | **N\A** | **N\A** | **N\A** |
| **Kuckelman et al.** | **6** | **N\A** | **N\A** |
| **Jeblick et.al** | **N\A** | **N\A** | **N\A** |
| **Scheschenja et al.** | **1** | **N\A** | **N\A** |
| **Gordon et al.** | **N\A** | **N\A** | **N\A** |
| **Schmidt et al.** | **4** | **N\A** | **N\A** |

**Table 5**. A summary of AI negative performance

| **Study** | **Incorrect Information** | **Missing Information** | **Inaccurate Information** | **Potentially Harmfull Information** |
| --- | --- | --- | --- | --- |
| **Lyu et al.** | **2 found in 10 reports** | **37 found in 10 reports** | **62 found in 10 reports** | **N/A** |
| **Li et al.** | **N/A** | **N/A** | **N/A** | **N/A** |
| **Grewal et al.** | **N/A** | **N/A** | **N/A** | **N/A** |
| **Kuckelman et al.** | **N/A** | **Explanation about bone biopsy did not include possible damage to adjacent structures that may occur during biopsy** | **N/A** | **N/A** |
| **Jeblick et.al** | **23 out of 45 reports (51%)** | **10 out of 45 reports (22%)** | **N/A** | **16 out of reports 45 (36%)** |
| **Scheschenja et al.** | **Chat-GPT 3.5: 5.3% Chat-GPT 4 : 2.3%** | **N/A** | **N/A** | **N/A** |
| **Gordon et al.** | **N/A** | **N/A** | **N/A** | **N/A** |
| **Schmidt et al.** | **N/A** | **7 out of 13 reports (53.8%)** | **N/A** | **3 out of 13 reports  (23%)** |

**Table 6**. Presents examples of text simplification performed by LLMs

| **Study** | **Shortening** | **Explaining Medical Jargon** | **Integration** | **Suggestions** |
| --- | --- | --- | --- | --- |
| **Lyu et al.** | **“The pleura, heart and blood vessels are normal, and there is no sign of cancer in the lymph nodes.”** | **“There is a small 1 mm area in the right middle lobe that looks like a granuloma, which is a small area of inflammation that is usually not concerning.”** | **“There is also a 6 mm granuloma in the right lower lobe, but it has not changed since a previous CT scan done in August 2021.”** | **“Follow up with doctors”** |
| **Li et al.** | **"There are no other significant findings in the rest of the body"** | **"The report mentions that your liver has a condition called "hepatic steatosis", which means there is excess fat in the liver."** | **"Your spleen is slightly larger than normal, but this has not changed since the last time you had a CT scan"** | **N/A** |
| **Grewal et al.** | **N/A** | **"5 cm arterially enhancing lesion": A lesion (abnormal tissue) measuring 5 centimetres is seen in the liver** | **N/A** | **"...you should consult your doctor to discuss these findings, receive a proper diagnosis, and discuss any necessary next steps.”** |
| **Kuckelman et al.** | **N/A** | **CT abdomen is a type of specialized X-ray that helps doctors see the organs, blood vessels, and bones in your abdomen** | **N/A** | **talk to your doctor so you get properly prepared for your MRI** |
| **Jeblick et.al** | **N/A** | **N/A** | **N/A** | **N/A** |
| **Scheschenja et al.** | **N/A** | **N/A** | **N/A** | **N/A** |
| **Gordon et al.** | **N/A** | **N/A** | **N/A** | **N/A** |
| **Schmidt et al.** | **N/A** | **N/A** | **N/A** | **N/A** |

**Table 7**. Presents examples of prompt inputs for LLMs

| **Study** | **1st Prompt** | **Optimized Prompt** |
| --- | --- | --- |
| **Lyu et al.** | **1. Please translate a radiology report into plain language that is easy to understand.**  **2. Please provide some suggestions for the patient.**  **3. Please provide some suggestions for the healthcare provider.** | **Please help translate a radiology report into plain language in the following format:  First paragraph introduces screening description description including reason for screening, screening time, protocol, patient background, and comparison date;  Second paragraph talks about specific findings: how many nodules detected, each lung nodule’s precise position and size, findings on lungs, heart, pleura, coronary artery calcification, mediastinum/hilum/axilla, and other findings. Please don’t leave out any information about findings;  Third paragraph talks about conclusions, including overall lung-rads category, management recommendation and follow-up date, based on lesion;  If there are incidental findings, please introduce in the fourth paragraph.** |
| **Li et al.** | **“Explain this radiology report to a patient in layman's terms in second person: <Report Text>.”** | **N/A** |
| **Grewal et al.** | **Got my MRI results back today and my MRI abdomen report states ' 5 cm arterially enhancing lesion in segment VII of liver with arterial phase enhancement and washout on portal venous phase and a pseudocapsule, compatible with HCC (Liver Reporting & Data System (LI-RADS®))'. Please simplify this report for me.** | **N/A** |
| **Kuckelman et al.** | **"What is a CT of the abdomen"** | **N/A** |
| **Jeblick et.al** | **N/A** | **“Explain this medical report to a child using simple language:”** |
| **Scheschenja et al.** | **Please answer the following questions about percutaneous transluminal angioplasty in peripheral arterial disease.”** | **N/A** |
| **Gordon et al.** | **“Provide an accurate and easy-to understand response that is suited for an average person** | **N/A** |
| **Schmidt et al.** | **“Simplify the following finding”** | **“Explain the following MRI report of the knee joint in simple language:”** |
