## Supplemental Figures 1-3 for "Large language models in simplifying radiological reports: systematic review"

**Figure 1.** Flow diagram of the search and inclusion process in the study

**Identification of studies via databases and registers**

Records removed *before screening*:

Duplicate records removed (n = 0)

Records marked as ineligible by automation tools (n = 0)

Records removed for other reasons (n = 0)

Records identified from*:

Databases (n = 729)

Registers (n = 0)

**Identification**

Records screened

(n = 729)

Records excluded**

(n = 12)

Reports sought for retrieval

(n =717)

Reports not retrieved

(n =0)

**Screening**

Reports assessed for eligibility

(n =8)

Reports excluded:

Irrelevant (n =666)

Not original study 2 (n =41)

Non-English (n =2)

Studies included in review

(n =8)

Reports of included studies

(n = 8)

**Included**

**Figure 1**. Flow Diagram of the Inclusion Process. Flow diagram of the search and inclusion process based on the Preferred Reporting Items for Systematic Reviews and Meta-Analyses (PRISMA) guidelines, January 2024.

**Figure 2.** Risk of Bias and Applicability Judgments in tailored QUADAS-2

|  | **RISK OF BIAS** | | | |  | **APPLICABILITY CONCERNS** | | |
| --- | --- | --- | --- | --- | --- | --- | --- | --- |
|  | **PATIENT SELECTION** | **INDEX TEST** | **REFERENCE STANDARD** | **FLOW AND TIMING** |  | **PATIENT SELECTION** | **INDEX TEST** | **REFERENCE STANDARD** |
| Lyu et al. | **■** | **?** | **▲** | **■** |  | **■** | **■** | **■** |
| Li et al. | **■** | **■** | **?** | **■** |  | **▲** | **■** | **■** |
| Grewal et al. | **?** | **?** | **?** | **■** |  | **?** | **?** | **?** |
| Kuckelman et al. | **■** | **■** | **■** | **■** |  | **■** | **■** | **■** |
| Jeblick et.al | **■** | **▲** | **▲** | **■** |  | **■** | **■** | **■** |
| Scheschenja et al. | **▲** | **▲** | **■** | **■** |  | **■** | **■** | **■** |
| Gordon et al. | **■** | **■** | **■** | **■** |  | **■** | **■** | **■** |
| Schmidt et al. | **■** | **▲** | **■** | **■** |  | **■** | **■** | **■** |

| Low Risk | **■** |
| --- | --- |
| High Risk | **▲** |
| Unclear Risk | **?** |

**Figure 2:** QUADAS-2 table for potential bias and applicability. Risk of bias and applicability were evaluated using the tailored QUADAS-2 tool, January 2024.

**Figure 3**. Main attributes reviewed in the study.

|  | **Lyu et al.** | | | | |  |  | **Li et al.** | | | | |  |  | **Grewal et al.** | | | | |  |  | **Kuckelman et al.** |
| --- | --- | --- | --- | --- | --- | --- | --- | --- | --- | --- | --- | --- | --- | --- | --- | --- | --- | --- | --- | --- | --- | --- |
|  | **Jeblick et.al** | | | | |  |  | **Scheschenja et al.** | | | | |  |  | **Gordon et al.** | | | | |  |  | **Schmidt et al.** |

| **Legend:** | | |  |  |  |
| --- | --- | --- | --- | --- | --- |
|  |  |  |  |  | **Simplified radiological reports** |
|  |  |  |  |  | **Answered patient questions** |
|  |  |  |  |  | **Patient evaluation** |
|  |  |  |  |  | **Compared initial prompt to optimized prompt** |
|  |  |  |  |  | **Generated inadequate outcome** |

**Figure 3**. Presents a graphical illustration of main attributes reviewed in our study. The attributes are color coded and serve as indication for each study.
